# Psychological Readiness, Injury-Related Fear, and Persistent Knee Symptoms After Anterior Cruciate Ligament Reconstruction in Riyadh, Saudi Arabia: A Cross-Sectional Study

**DOI:** 10.1101/2025.05.14.25327357

**Authors:** Turki Fahid Alqahtani, Tariq Yousef Alhomoud, Mohammed Jafar Albin Ahmed, Faisal Naif Al-Mohaisen, Abbas Abdullah Alhejji, Abdulaziz Abdulrahman Alrabiah

**Affiliations:** Department of Anesthesia and Surgery, College of Medicine, Imam Mohammad Ibn Saud Islamic University (IMSIU), Riyadh, Saudi Arabia; College of Medicine, Imam Mohammad Ibn Saud Islamic University (IMSIU), Riyadh, Saudi Arabia; College of Medicine, King Saud University (KSU), Riyadh, Saudi Arabia

**Keywords:** ACL reconstruction, psychological readiness, kinesiophobia, ACL-RSI, TSK-11, return to sport, Saudi Arabia

## Abstract

**Background:** Anterior cruciate ligament (ACL) ruptures are highly prevalent among physically active individuals. Although patients often undergo successful surgical reconstruction, many experience psychological challenges that influence recovery and return to sport.

**Objective:** This study aimed to assess psychological readiness and injury-related fear among individuals who underwent ACL reconstruction.

**Study Design:** Cross-sectional study

**Methods:** We distributed a structured, self-administered electronic questionnaire to 96 participants in Riyadh, Saudi Arabia. We measured psychological readiness using the ACL Return to Sport after Injury (ACL-RSI) scale and assessed fear of reinjury using the Tampa Scale for Kinesiophobia-11 (TSK-11).

**Results:** Participants reported moderate psychological readiness (ACL-RSI: Mean = 47, SD = 18) and kinesiophobia (TSK-11: Mean = 27, SD = 6). Participants over 40 years old demonstrated significantly lower readiness scores and higher levels of kinesiophobia than younger groups.

**Conclusion:** Psychological readiness and injury-related fear are critical components of recovery after ACL reconstruction. Rehabilitation programs should address both physical and psychological dimensions to improve patient outcomes. Future research should explore the effectiveness of targeted psychological interventions in ACL recovery.

## Introduction

Anterior cruciate ligament (ACL) injuries are among the most common knee injuries, particularly in sports involving rapid deceleration, pivoting, or contact. Globally, the incidence of ACL injuries has been steadily increasing, with approximately 200,000 cases reported annually in the United States alone. In Saudi Arabia, recent data suggest that ACL injuries are highly prevalent among young, physically active individuals, especially football players, where rates have reached over 75% in some regional studies.

The standard treatment for ACL rupture, especially in athletes and young individuals, involves surgical reconstruction followed by a structured rehabilitation program. While physical outcomes such as knee stability and range of motion are well studied, growing evidence highlights the importance of psychological factors in predicting successful return to sport. Key psychological constructs—such as fear of reinjury, self-efficacy, and emotional readiness—are now considered vital predictors of functional recovery.

Instruments such as the ACL Return to Sport after Injury (ACL-RSI) scale and the Tampa Scale for Kinesiophobia (TSK-11) have been widely used to quantify emotional responses and fear avoidance behaviors after ACL reconstruction. These tools have been translated and validated in Arabic-speaking populations, offering an opportunity to investigate region-specific factors that may influence psychological outcomes.

Despite the increasing recognition of psychological readiness in sports medicine literature, few studies in the Middle East—particularly in Saudi Arabia—have comprehensively assessed these variables following ACL reconstruction. This study aimed to evaluate psychological readiness and injury-related fear among patients in Riyadh, using validated scales, and to explore associations with age, sex, injury mechanism, and rehabilitation engagement.

Anterior cruciate ligament (ACL) injuries frequently affect physically active individuals, often requiring surgical intervention to restore knee stability and functional mobility. While surgical reconstruction and physical rehabilitation remain standard components of ACL injury management, psychological recovery increasingly defines long-term outcomes, particularly the return to pre-injury activity levels [1,2].

## Methods

We conducted a cross-sectional study in Riyadh, Saudi Arabia, targeting individuals who had undergone anterior cruciate ligament (ACL) reconstruction within the past five years. We included participants aged 18 years or older who were capable of completing an Arabic-language survey. We excluded individuals with cognitive or psychiatric impairments that could interfere with accurate questionnaire completion.

We developed a structured, self-administered questionnaire based on validated Arabic versions of the ACL Return to Sport after Injury (ACL-RSI) scale and the Tampa Scale for Kinesiophobia-11 (TSK-11). The survey included four domains: (1) demographic information, (2) injury and surgical details, (3) rehabilitation status, and (4) psychological readiness. The ACL-RSI scale comprises items evaluating confidence, risk appraisal, and emotional readiness on a Likert scale. The TSK-11 measures fear of movement and reinjury, also using a Likert scale. Both instruments have demonstrated strong reliability and construct validity in prior research.

We distributed the electronic survey using convenience sampling via social media platforms (Twitter, WhatsApp, and Telegram) and collected responses between [insert start and end dates]. We calculated a sample size of 96 using a 95% confidence interval, 10% margin of error, and an assumed prevalence of 50% for psychological readiness.

We analyzed the data using IBM SPSS version 29.0. Categorical variables were summarized as frequencies and percentages, while continuous variables were presented as means and standard deviations or medians with interquartile ranges, depending on distribution. We used t-tests and ANOVA to compare mean ACL-RSI and TSK-11 scores across subgroups. We assessed relationships using Pearson correlation and conducted multiple regression analyses to identify independent predictors of psychological readiness. Statistical significance was set at p < 0.05.

We obtained ethical approval from the Institutional Review Board of Al-Imam Muhammad Ibn Saud Islamic University. All participants provided informed electronic consent before participation.

We conducted a cross-sectional study in Riyadh, Saudi Arabia, targeting individuals who had undergone anterior cruciate ligament (ACL) reconstruction. Eligible participants were at least 18 years old and capable of completing an Arabic-language questionnaire. We excluded individuals with cognitive impairments that could interfere with survey completion.

## Results

From 112 potential participants, 108 agreed to participate in the study (96.4% response rate), with 74 having ACL injuries and 34 without injuries. The study population was predominantly male (n=107, 99.1%), with only one female participant (n=1, 0.9%). Age distribution showed that most participants were between 26-29 years (n=39, 36.1%), followed by 22-25 years (n=32, 29.6%). Nearly two-thirds of participants were single or divorced (n=70, 64.8%), and half resided in the Central Region (n=55, 50.9%).

When analyzing differences between injured and non-injured groups, age distribution showed statistically significant variations (p<0.001). Most notably, the 26-29 age group had a substantially higher proportion of ACL injuries (n=37, 50.0%) compared to non-injuries (n=2, 5.9%). Conversely, those aged over 40 showed lower injury rates (n=4, 5.4%) despite higher representation in the non-injured group (n=12, 35.3%). Other demographic variables, including sex (p=0.315), marital status (p=0.088), and place of residence (p=0.059), did not show statistically significant differences between the groups. **(Table 1)**

**Table 1.** Demographic Characteristics and ACL Injury Status of Study Participants.

| Variable | No N (%) | Yes N (%) | Total N (%) | p-Value |
| --- | --- | --- | --- | --- |
| Sex |  |  |  |  |
| Female | 1 (2.9%) | 0 (0.0%) | 1 (0.9%) | 0.315 |
| Male | 33 (97.1%) | 74 (100.0%) | 107 (99.1%) |  |
| Age |  |  |  |  |
| 18-21 | 6 (17.6%) | 7 (9.5%) | 13 (12.0%) | <0.001 |
| 22-25 | 12 (35.3%) | 20 (27.0%) | 32 (29.6%) |  |
| 26-29 | 2 (5.9%) | 37 (50.0%) | 39 (36.1%) |  |
| 30-39 | 2 (5.9%) | 6 (8.1%) | 8 (7.4%) |  |
| >40 | 12 (35.3%) | 4 (5.4%) | 16 (14.8%) |  |
| Marital Status |  |  |  |  |
| Single/Divorced | 18 (52.9%) | 52 (70.3%) | 70 (64.8%) | 0.088 |
| Married | 16 (47.1%) | 22 (29.7%) | 38 (35.2%) |  |
| Place of Residence |  |  |  |  |
| Central Region | 20 (58.8%) | 35 (47.3%) | 55 (50.9%) | 0.059 |
| North/East/West | 0 (0.0%) | 10 (13.5%) | 10 (9.3%) |  |
| Southern Region | 14 (41.2%) | 29 (39.2%) | 43 (39.8%) |  |

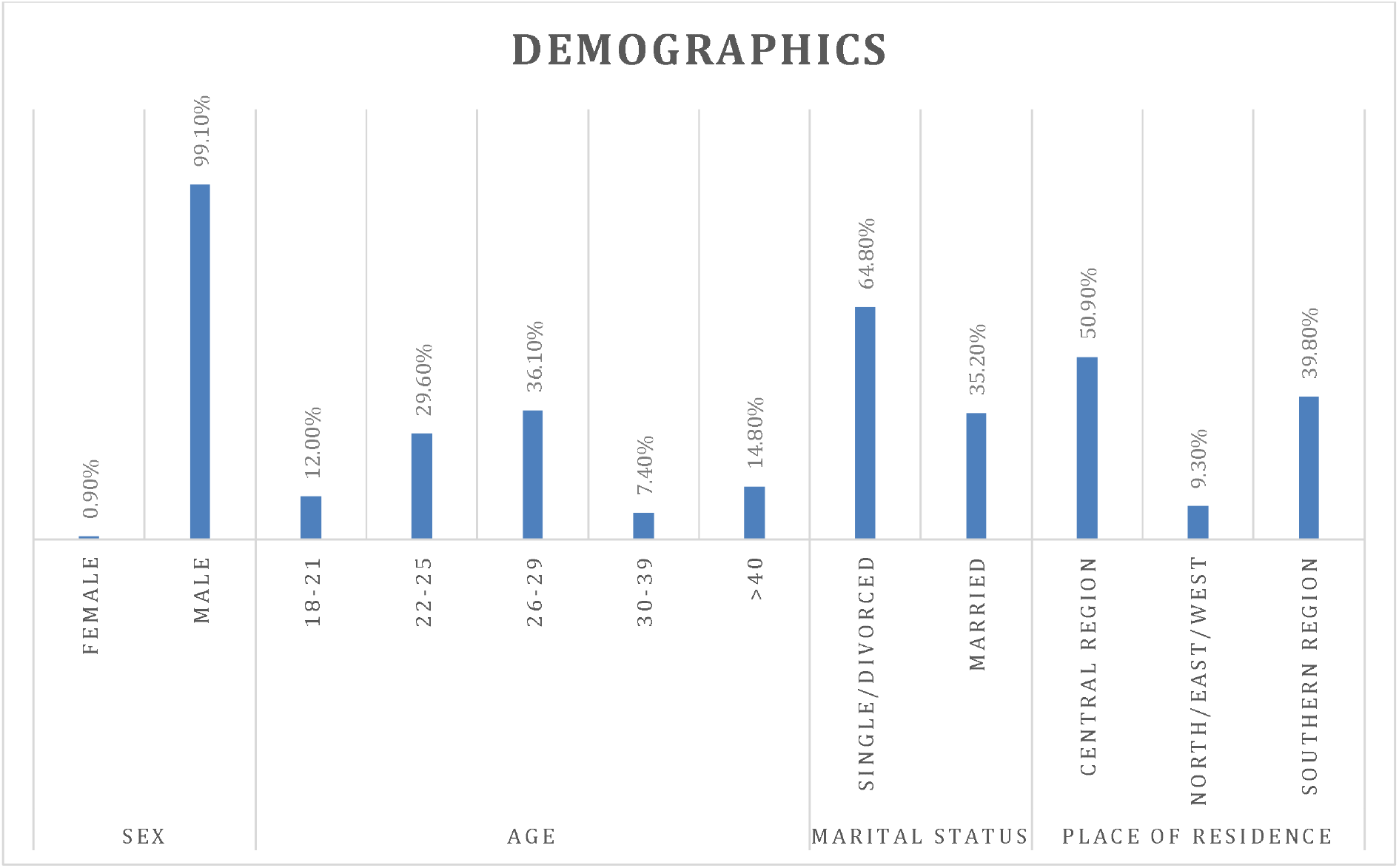

Among the 74 participants with ACL injuries, the majority received physical therapy (n=61, 82.4%), while a smaller proportion did not undergo physical therapy (n=13, 17.6%). Regarding ACL reconstructions, most participants underwent single reconstruction surgery (n=47, 63.5%), while some had not undergone any reconstruction (n=23, 31.1%), and a small number required double reconstruction (n=4, 5.4%).

For those who underwent ACL reconstruction (n=50 available), the hamstring tendon was the predominant graft choice (n=41, 82%), followed by quadriceps tendon (n=6, 12%), and patellar tendon (n=3, 6%). **(Table 2)**

**Table 2.** Treatment Characteristics of ACL-Injured Participants.

| Variable | N (%) |
| --- | --- |
| **Did you have physical therapy for the injury?** |  |
| No | 13 (17.6%) |
| Yes | 61 (82.4%) |
| **How many ACL reconstructions have you had?** |  |
| 0 | 23 (31.1%) |
| 1 | 47 (63.5%) |
| 2 | 4 (5.4%) |
| **Where did you get the patch for the ligament?** |  |
| Hamstring Tendon | 41 (82%) |
| Patellar Tendon (Kneecap) | 3 (6%) |
| Quadriceps Tendon (Anterior Thigh) | 6 (12%) |

The ACL Return to Sport after Injury (ACL-RSI) scale and its components were assessed on a scale of 0-10, with higher scores indicating better psychological readiness. The overall ACL-RSI score averaged 47 (SD=18) with a range of 11-92.

Participants showed the highest confidence in feeling relaxed during exercise (Mean=6, SD=2), while demonstrating moderate levels (Mean=5, SD=3) across several domains including confidence in returning to pre-injury performance levels, general exercise concerns, and sports performance ability. The lowest mean scores (Mean=4, SD=2-3) were observed in areas related to re-injury concerns, knee stability under pressure, and surgical/rehabilitation concerns. **(Table 3)**

**Table 3.** Psychological Readiness and Fear Assessment Scores in ACL-Injured Participants.

| Variable | Mean (SD) | Range |
| --- | --- | --- |
| **Are you confident that you can return to playing at the same level as before the injury?** | 5 (3) | 0 – 10 |
| **Do you think you are likely to injure your knee again when you play again?** | 4 (2) | 0 – 10 |
| **Are you worried about exercising?** | 5 (3) | 0 – 10 |
| **Are you confident that you can play your sport without worrying about your knees?** | 5 (3) | 0 – 10 |
| **Are you confident about returning to sports without worrying about your knee?** | 5 (2) | 0 – 10 |
| **Are you frustrated that you have to think about your knee when playing your sport?** | 5 (3) | 0 – 10 |
| **Are you afraid of re-injuring your knee due to your sport?** | 4 (3) | 0 – 10 |
| **Are you confident that your knee can hold up under pressure?** | 4 (2) | 0 – 10 |
| **Are you afraid of accidentally injuring your knee while playing your sport?** | 4 (3) | 0 – 10 |
| **Does the thought of surgery and rehabilitation keep you from playing your sport?** | 4 (3) | 0 – 10 |
| **Are you confident in your ability to perform well in sports?** | 5 (3) | 0 – 10 |
| **Do you feel relaxed when you exercise?** | 6 (2) | 0 – 10 |
| **ACL-RSI** | 47 (18) | 11 – 92 |

The TSK-11 assessment yielded an overall mean score of 27 (SD=6, range=13-44), indicating moderate levels of kinesiophobia in the study population. The TSK-11 uses a 4-point Likert scale where 1 represents “Strongly Disagree” and 4 represents “Strongly Agree”.

Participants showed highest concerns (Mean=3, SD=1) in areas related to exercise-induced injury fear, pain increase with activity, pain as a warning signal, and exercise during pain. Lower scores (Mean=2, SD=1) were reported for items related to bodily damage perception, medical condition recognition, and long-term injury risk. **(Table 4)**

**Table 4.**
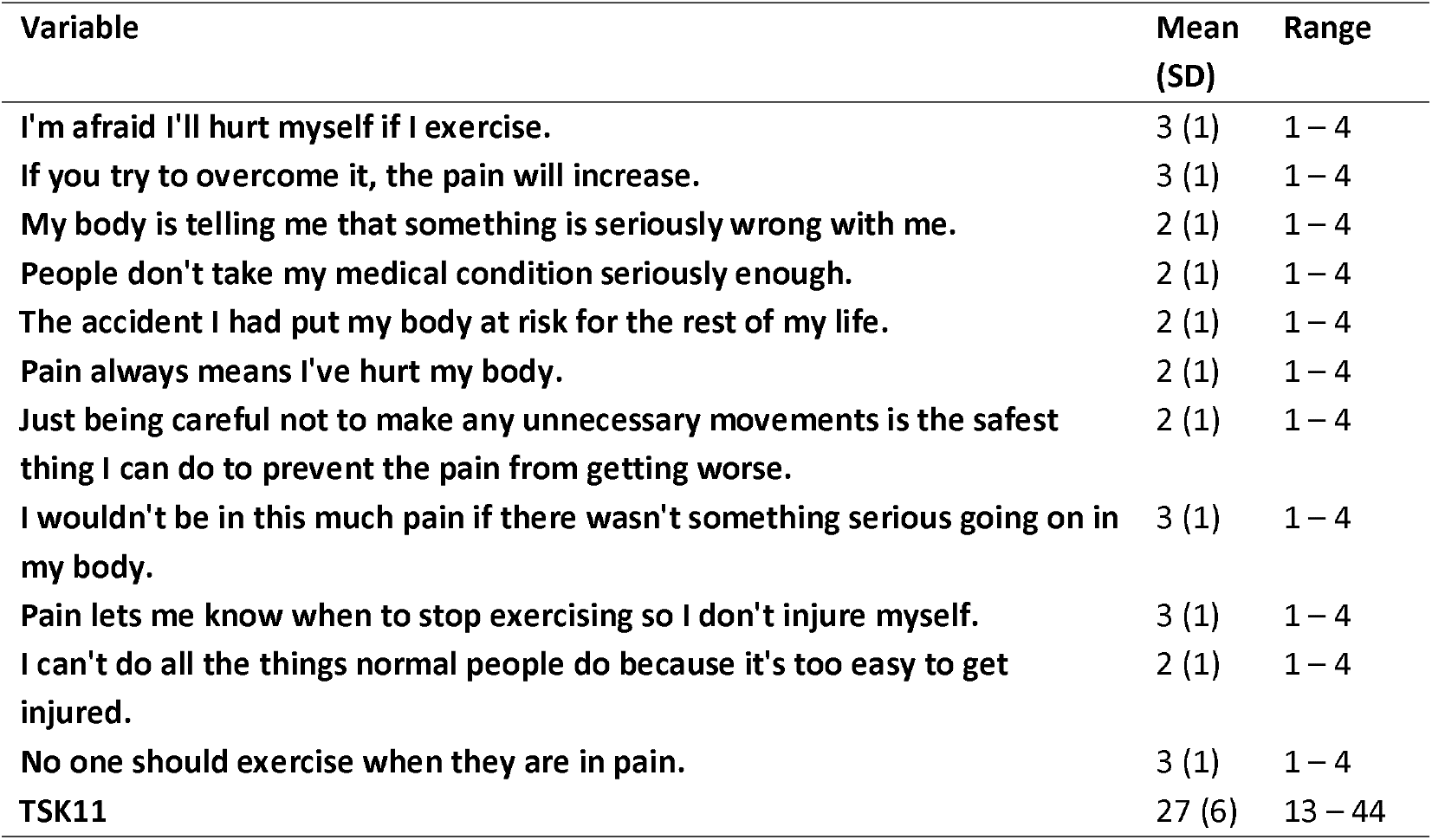
Tampa Scale for Kinesiophobia-11 (TSK-11) Assessment Results.

Age significantly influenced both psychological readiness (ACL-RSI) (p=0.042) and kinesiophobia (TSK-11) scores (p=0.026). Participants aged >40 years demonstrated the lowest psychological readiness (Mean=26.9, SD=18.0) and highest kinesiophobia (Mean=36.3, SD=1.5), while those aged 30-39 showed higher psychological readiness (Mean=52.9, SD=26.4) and lower kinesiophobia (Mean=23.7, SD=6.7).

Marital status showed a significant difference in TSK-11 scores (p=0.041), with married participants displaying higher kinesiophobia (Mean=29.0, SD=5.8) compared to single/divorced individuals (Mean=25.8, SD=6.4). However, ACL-RSI scores did not significantly differ by marital status (p=0.247).

Geographic location and physical therapy intervention did not significantly impact either psychological readiness or kinesiophobia (p>0.05). However, those who received physical therapy showed a trend toward lower kinesiophobia (Mean=26.0, SD=5.8) compared to those who did not (Mean=30.3, SD=7.8). **(Table 5)**

**Table 5.** Psychological Readiness and Kinesiophobia Scores by Demographic and Treatment.

| Variable | ACL-RSI Mean (SD) | p-Value | TSK-11 Mean (SD) | p-Value |
| --- | --- | --- | --- | --- |
| **Age** |  |  |  |  |
| 18-21 | 53.6 (16.5) | 0.042 | 26.0 (4.8) | 0.026 |
| 22-25 | 41.0 (19.0) |  | 26.7 (6.7) |  |
| 26-29 | 49.8 (14.7) |  | 26.4 (5.9) |  |
| 30-39 | 52.9 (26.4) |  | 23.7 (6.7) |  |
| >40 | 26.9 (18.0) |  | 36.3 (1.5) |  |
| **Marital Status** |  |  |  |  |
| Single/Divorced | 48.3 (18.6) | 0.247 | 25.8 (6.4) | 0.041 |
| Married | 43.1 (16.8) |  | 29.0 (5.8) |  |
| **Place of Residence in the Kingdom** |  |  |  |  |
| Central Region | 45.2 (16.7) | 0.115 | 26.5 (6.9) | 0.317 |
| North/East/West | 57.8 (8.6) |  | 24.4 (4.3) |  |
| Southern Region | 44.9 (21.0) |  | 27.9 (6.1) |  |
| **Did you have physical therapy?** |  |  |  |  |
| No | 43.8 (12.0) | 0.398 | 30.3 (7.8) | 0.079 |
| Yes | 47.4 (19.2) |  | 26.0 (5.8) |  |

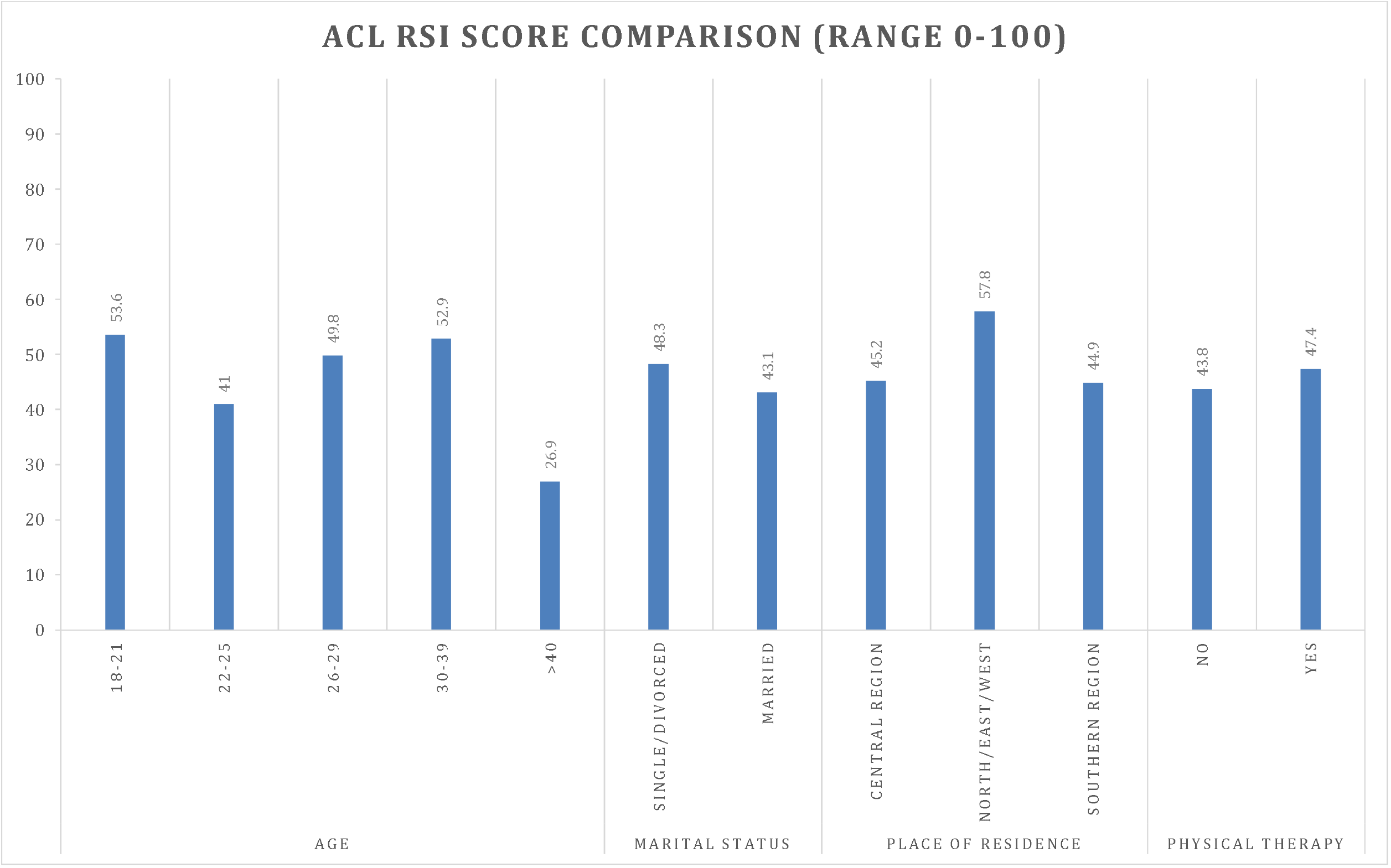

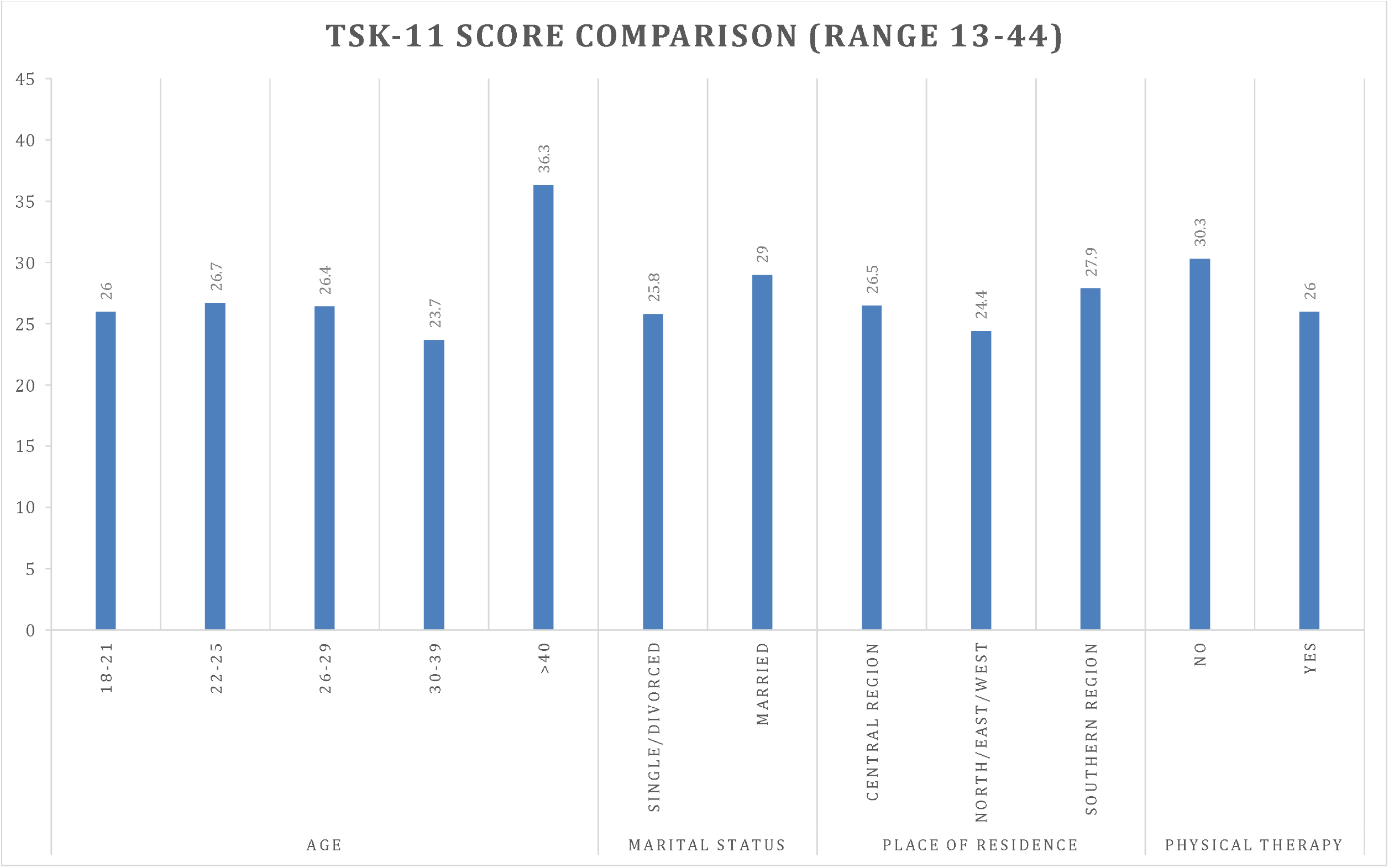

Analysis revealed significant correlations between the study variables. A strong negative correlation was observed between ACL-RSI and TSK-11 scores (r=-0.485, p<0.001), indicating that higher psychological readiness was associated with lower kinesiophobia.

The number of ACL reconstructions showed a significant negative correlation with TSK-11 scores (r=-0.278, p=0.017), suggesting that participants with more reconstructions tended to have lower kinesiophobia levels. However, there was no significant correlation between the number of ACL reconstructions and ACL-RSI scores (r=0.099, p=0.400).

These findings suggest that while multiple reconstructions might lead to reduced movement-related fear, they don’t necessarily translate to improved psychological readiness for return to sport. (Table 6)

**Table 6.** Correlations Between Number of ACL Reconstructions, Psychological Readiness, and Kinesiophobia.

|  |  | No. of ACL reconstructions | ACL-RSI | TSK11 |
| --- | --- | --- | --- | --- |
| No. of ACL reconstructions | Pearson Correlation | 1 | .099 | -.278* |
|  | Sig. (2-tailed) |  | .400 | .017 |
| ACL-RSI | Pearson Correlation | .099 | 1 | -.485** |
|  | Sig. (2-tailed) | .400 |  | <.001 |
| TSK11 | Pearson Correlation | -.278* | -.485** | 1 |
|  | Sig. (2-tailed) | .017 | <.001 |  |

\*. Correlation is significant at the 0.05 level (2-tailed). \*\*. Correlation is significant at the 0.01 level (2-tailed).

The regression analysis revealed that none of the predictors reached statistical significance at p<0.05, though some variables approached significance. Age showed trending relationships with ACL-RSI scores, particularly for the 22-25 age group (B=-15.72, β=-.39, p=.054) and those >40 years (B=-22.02, β=-.28, p=.058), suggesting a tendency toward lower psychological readiness in these age groups compared to the 18-21 reference group.

Geographic location showed a marginal effect for the North/East/West region (B=11.62, β=.22, p=.080) compared to the Central Region, indicating a potential trend toward higher psychological readiness in these regions. Other variables, including marital status (B=-7.00, β=-.18, p=.181), physical therapy (B=0.18, β=.00, p=.975), and number of ACL reconstructions (B=4.29, β=.13, p=.299), did not demonstrate significant associations with ACL-RSI scores. (Table 7)

**Table 7.** Regression Analysis Predicting ACL-RSI Scores.

| Predictor | B | $\beta$ (Standardized) | p-Value | 95% CI for B |
| --- | --- | --- | --- | --- |
| Intercept (Constant) | 51.50 | — | < .001 | [34.75, 68.25] |
| Age (Reference: 18-21) |  |  |  |  |
| 22-25 | -15.72 | -.39 | .054 | [-31.69, 0.26] |
| 26-29 | -5.84 | -.16 | .424 | [-20.32, 8.64] |
| 30-39 | -3.67 | -.06 | .712 | [-23.49, 16.14] |
| >40 | -22.02 | -.28 | .058 | [-44.82, 0.77] |
| **Marital Status (Reference: Single/Divorced)** |  |  |  |  |
| Married | -7.00 | -.18 | .181 | [-17.36, 3.35] |
| **Place of Residence (Reference: Central Region)** |  |  |  |  |
| North/East/West | 11.62 | .22 | .080 | [-1.41, 24.65] |
| Southern Region | 2.83 | .08 | .534 | [-6.22, 11.88] |
| **Physical Therapy (Yes vs. No)** | 0.18 | .00 | .975 | [-11.09, 11.45] |
| **Number of ACL Reconstructions** | 4.29 | .13 | .299 | [-3.89, 12.48] |

*B* = unstandardized coefficient; $\beta$ = standardized coefficient; CI = confidence interval.

Age >40 years showed a trending positive relationship with kinesiophobia (B=7.50, β=.27, p=.053), suggesting older participants tended to have higher kinesiophobia scores. Married status also showed a marginal positive association (B=3.04, β=.22, p=.083), indicating a tendency toward higher kinesiophobia compared to single/divorced participants.

The number of ACL reconstructions showed a trending negative relationship with kinesiophobia (B=-2.55, β=-.22, p=.066), suggesting that more reconstructions might be associated with lower fear levels. Physical therapy showed a non-significant negative relationship (B=-2.75, β=-.17, p=.148), while geographic location demonstrated no significant associations with TSK-11 scores. (Table 8)

**Table 8.** Regression Analysis Predicting TSK-11 Scores.

| Predictor | B | $\beta$ (Standardized) | p-Value | 95% CI for B |
| --- | --- | --- | --- | --- |
| **Intercept (Constant)** | 28.50 | — | < .001 | [22.93, 34.07] |
| **Age (Reference: 18-21)** |  |  |  |  |
| 22-25 | 2.25 | .16 | .400 | [-3.06, 7.57] |
| 26-29 | 1.04 | .08 | .668 | [-3.78, 5.85] |
| 30-39 | -1.46 | -.06 | .660 | [-8.05, 5.13] |
| >40 | 7.50 | .27 | .053 | [-0.08, 15.08] |
| **Marital Status (Reference: Single/Divorced)** |  |  |  |  |
| Married | 3.04 | .22 | .083 | [-0.41, 6.48] |
| **Place of Residence (Reference: Central Region)** |  |  |  |  |
| North/East/West | -1.41 | -.08 | .518 | [-5.74, 2.92] |
| Southern Region | 0.73 | .06 | .629 | [-2.28, 3.74] |
| **Physical Therapy (Yes vs. No)** | -2.75 | -.17 | .148 | [-6.50, 1.00] |
| **Number of ACL Reconstructions** | -2.55 | -.22 | .066 | [-5.27, 0.18] |

$B$ = unstandardized coefficient; $\beta$ = standardized coefficient; CI = confidence interval.

## Conclusion

The study revealed moderate levels of psychological readiness (ACL-RSI: Mean=47, SD=18) and kinesiophobia (TSK-11: Mean=27, SD=6) among ACL-injured patients in Riyadh, Saudi Arabia. Age emerged as a significant factor, with participants over 40 showing lower psychological readiness and higher kinesiophobia. A strong negative correlation was found between psychological readiness and kinesiophobia (r=-0.485, p<0.001). While most participants received physical therapy (82.4%) and underwent single ACL reconstruction (63.5%), these interventions showed limited impact on psychological outcomes. The findings suggest that psychological rehabilitation should be age-specific and integrated into the standard ACL injury treatment protocol, particularly focusing on older patients who demonstrate higher kinesiophobia and lower return-to-sport confidence.

## Discussion

This study examined psychological readiness and injury-related fear among individuals who underwent ACL reconstruction in Riyadh, Saudi Arabia. The findings revealed moderate psychological readiness and elevated kinesiophobia levels, highlighting the need for psychological support during rehabilitation.

Participants over the age of 40 exhibited lower ACL-RSI scores and higher levels of kinesiophobia compared to younger individuals. This aligns with previous research indicating that older patients may experience greater psychological distress due to longer recovery times, reduced physical conditioning, or increased reinjury concerns [7,8]. Age-specific psychological interventions could enhance recovery outcomes in this population.

Interestingly, participants who sustained injuries outside sports contexts reported higher psychological readiness. This may reflect lower perceived performance pressure or reduced expectations of athletic return compared to sport-related injuries, where competitive drive and performance goals often amplify psychological burden [9].

Contrary to expectations, participants who did not attend regular physical therapy sessions had higher ACL-RSI scores. One explanation could be that individuals with higher initial self-efficacy or milder injury severity felt less dependent on structured rehabilitation programs. Alternatively, this finding could suggest a gap in motivation, adherence, or access to psychological resources among those in therapy—warranting further investigation [10].

Sex, height, weight, time since surgery, and graft type showed no significant association with psychological readiness, supporting prior studies emphasizing the importance of emotional and perceptual variables over physical or procedural metrics [11,12].

These results reinforce the value of integrating psychological assessments into postoperative care. Instruments like ACL-RSI and TSK-11 allow clinicians to identify patients at risk for poor recovery trajectories due to emotional barriers. Tailored psychological interventions, including cognitive behavioral therapy or return-to-sport counseling, could optimize outcomes.

Future research should adopt longitudinal designs to track psychological adaptation over time and evaluate the efficacy of multidisciplinary rehabilitation strategies. Expanding this research within Saudi Arabia and other Gulf countries could help establish culturally responsive approaches to recovery.

Participants reported moderate psychological readiness to return to sport. The average ACL-RSI score was 47 (SD = 18), while the average TSK-11 score was 27 (SD = 6). Older participants (>40 years) showed significantly lower ACL-RSI scores and higher kinesiophobia scores than their younger counterparts (p < 0.05).

The analysis revealed no statistically significant difference in psychological readiness based on sex or time since surgery. However, participants who sustained injuries in non-sport-related incidents demonstrated significantly higher readiness levels compared to those with sports-related injuries (p = 0.038). Additionally, participants who did not undergo regular physical therapy reported significantly higher ACL-RSI scores than those who did (p = 0.021).

Correlation analysis showed no significant relationship between psychological readiness scores and physical attributes such as height (r = 0.056, p = 0.463), weight (r = 0.028, p = 0.713), or the time between injury and surgery (r = 0.091, p = 0.237).

Overall, these results suggest that psychological readiness depends more on psychosocial factors than on physical or procedural variables.

## Limitations

This study had several limitations. The cross-sectional design limited the ability to assess causality or psychological changes over time. The reliance on self-reported questionnaires may have introduced recall or response bias. Additionally, the sample primarily consisted of male participants, which restricts the generalizability of findings to females. Finally, the use of convenience sampling through online platforms may have excluded certain demographics, particularly individuals without internet access or familiarity with digital tools.

## Conclusion

Psychological readiness and injury-related fear significantly influence rehabilitation outcomes following ACL reconstruction. This study revealed moderate psychological readiness and elevated kinesiophobia among patients in Riyadh, with age and physical therapy adherence emerging as significant factors. These findings underscore the importance of integrating psychological assessment and support into ACL rehabilitation programs. Future research should adopt longitudinal designs and include broader, more diverse samples to further elucidate psychological recovery trajectories and inform individualized intervention strategies.

## Data Availability

All data produced in the present work are contained in the manuscript

## Conflict of Interest

The authors declare no conflicts of interest related to this study. Funding Statement: This research received no external funding.

## Ethical Approval

This study was approved by the Institutional Review Board at Imam Mohammad Ibn Saud Islamic University. Informed consent was obtained from all participants.

## Acknowledgments

The authors would like to thank all study participants for their time and cooperation.

## Notes

### Competing Interest Statement

The authors have declared no competing interest.

### Funding Statement

This study did not receive any funding

### Author Declarations

This study was approved by the Institutional Review Board at Imam Mohammad Ibn Saud Islamic University.

## References

1. Ardern CL, Taylor NF, Feller JA, Webster KE. A systematic review of the psychological factors associated with returning to sport following injury. Br J Sports Med. 2013;47(17):1120–6.

2. Ardern CL, Webster KE, Taylor NF, Feller JA. Return to sport following anterior cruciate ligament reconstruction surgery: a systematic review and meta-analysis of the state of play. Br J Sports Med. 2011;45(7):596–606.

3. Baez S, Harkey M, Birchmeier T, Triplett A, Collins K, Kuenze C. Psychological readiness, injury-related fear, and persistent knee symptoms after anterior cruciate ligament reconstruction. J Athl Train. 2023;58(11–12):998–1003.

4. Feller JA, Webster KE. Return to sport following anterior cruciate ligament reconstruction. Int Orthop. 2013;37(2):285–90.

5. Webster KE, Feller JA, Lambros C. Development and preliminary validation of a scale to measure the psychological impact of returning to sport following anterior cruciate ligament reconstruction surgery. Phys Ther Sport. 2008;9(1):9–15.

6. Woby SR, Roach NK, Urmston M, Watson PJ. Psychometric properties of the TSK-11: a shortened version of the Tampa Scale for Kinesiophobia. Pain. 2005;117(1–2):137–44.

7. Stigert M, Ashnai F, Thomeé R, Senorski EH, Beischer S. Physical inactivity 5–8 years after ACL reconstruction is associated with knee-related self-efficacy and psychological readiness to return to sport. BMJ Open Sport Exerc Med. 2023;9(4):e001687.

8. Ueda Y, Matsushita T, Shibata Y, Takiguchi K, Ono K, Kida A, et al. Association between meeting return-to-sport criteria and psychological readiness to return to sport after ACL reconstruction. Orthop J Sports Med. 2022;10(5):23259671221093985.

9. Presley J, Bailey L, Maloney K, Duncan B, Reid M, Juneau C, et al. The influence of mode-of-injury on psychological readiness for return-to-sport following ACL reconstruction: a matched-controlled study. Int J Sports Phys Ther. 2021;16(1):177–84.

10. Faleide AGH, Magnussen LH, Strand T, Bogen BE, Moe-Nilssen R, Mo IF, et al. The role of psychological readiness in return to sport assessment after ACL reconstruction. Am J Sports Med. 2021;49(5):1236–43.

11. Cronström A, Häger CK, Thorborg K, Ageberg E. Factors associated with sports function and psychological readiness to return to sports at 12 months after ACL reconstruction: a cross-sectional study. Am J Sports Med. 2023;51(12):3112–20.

12. Aizawa J, Hirohata K, Ohji S, Ohmi T, Mitomo S, Koga H, Yagishita K. Cross-sectional study on relationships between physical function and psychological readiness to return to sport after ACL reconstruction. BMC Sports Sci Med Rehabil. 2022;14(1):97.

